# Structural Brain Pathways Linking White Matter Hyperintensities to Pain Sensitivity

**DOI:** 10.64898/2026.07.14.26358028

**Authors:** Xianjing Liu, Torgil Riise Vangberg, Lieke M. Kuiper, Meike W. Vernooij, Audun Stubhaug, Ólöf Anna Steingrímsdóttir, Christian Magnus Page, Christopher Sivert Nielsen, Joyce B. J. van Meurs, Gennady V. Roshchupkin

## Abstract

People differ widely in their sensitivity to pain, and this variability is clinically relevant, yet the underlying structural brain mechanisms remain poorly understood. White matter hyperintensities (WMH), a common imaging marker of cerebral small vessel disease, are associated with microstructural abnormalities in white matter tracts and have also been linked to pain-related outcomes; however, the mechanisms linking WMH to altered pain perception remain unclear. We investigated whether WMH are linked to pain sensitivity through tract-specific microstructural alterations and cortical structural differences.

We analysed data from 1,448 participants (mean age 73 years; 53% women) in the population-based Rotterdam Study and independently replicated the findings in 1,522 participants (mean age 63 years; 52% women) from the population-based Tromsø Study. Pain sensitivity was quantified using the cold pressor test. Multimodal magnetic resonance imaging, including T1-weighted, fluid-attenuated inversion recovery and diffusion tensor imaging, was used to map WMH to predefined white matter tracts, derive tract-specific fractional anisotropy (FA), and estimate cortical measurements. Cox proportional hazards models assessed associations with pain sensitivity, and tract-specific mediation analyses evaluated whether white matter microstructure or tract-connected cortical regions mediated the relationship between white matter hyperintensities and pain sensitivity.

WMH were present in 20 of 27 predefined tracts and were associated with reduced FA in 18 tracts. Higher WMH burden was associated with greater pain sensitivity, particularly in the left anterior thalamic radiation and left superior thalamic radiation, while lower FA in the anterior thalamic radiation, medial lemniscus, superior thalamic radiation and inferior fronto-occipital fasciculus was associated with greater pain sensitivity. Mediation analyses showed that white matter microstructural disruption was the principal pathway linking WMH to pain sensitivity, with the strongest indirect effects observed through the inferior fronto-occipital fasciculus (44.6% mediated) and anterior thalamic radiation (32.6% mediated). Cortical atrophy in the precentral and postcentral gyri provided a smaller secondary pathway, mediating approximately 3–6% of the association between corticospinal or superior thalamic radiation WMH and pain sensitivity. Replication analyses supported these cortical mediation pathways, and meta-analysis strengthened the tract-specific associations.

Together, the results suggest that vascular white matter injury is associated with pain perception through specific structural pathways, with DTI-based markers appearing particularly sensitive to these relationships.

## Introduction

Pain sensitivity differs substantially across individuals, with marked variation in pain reporting, threshold, and tolerance when exposed to identical stimuli.^1^ This variability has important clinical implications: low pain sensitivity may contribute to under-recognition of serious conditions such as myocardial infarction,^2^ whereas high sensitivity is associated with increased risk of developing chronic pain,^3,4,5^ greater reliance on analgesics,^6^ and poorer recovery outcomes after injury.^7^ However, the neurobiological mechanisms underlying inter-individual differences in pain sensitivity remain incompletely understood. Accumulating evidence points to the central nervous system, particularly the brain, as a key driver of individual differences in pain perception.^8,9,10,11^ Uncovering structural brain features that underlie pain perception could advance our understanding of the mechanisms underlying individual differences in pain sensitivity and support earlier identification of high-risk individuals and the development of more targeted and effective interventions.

Several lines of evidence support that both gray and white matter brain structures are link to pain processing. Functional MRI studies have shown that cortical regions such as the insular cortex and postcentral gyrus are involved in sensory discrimination, interoception, and affective-motivational aspects of pain,^12,13^ while structural MRI studies have shown that changes in these areas have been associated with altered pain sensitivity.^14^ These cortical regions are interconnected through white matter pathways that enable the integration and transmission of information relevant to pain processing. Diffusion tensor imaging (DTI) studies provide evidence for the involvement of white matter in pain processing, with findings of increased mean diffusivity in the anterior thalamic radiation (ATR) in migraine patients and reduced fractional anisotropy in the same tract among individuals with interstitial cystitis/bladder pain syndrome.^15,16^

Initially, research on brain–pain relationships focused on identifying specific regions involved in pain processing, giving rise to the concept of a “pain matrix”.^17,18^ However, this notion of a fixed set of pain-processing regions has been challenged.^19^ Pain perception is increasingly viewed as a distributed and dynamic process involving interactions across widespread brain regions, prompting a shift in research toward characterizing whole-brain network properties and functional connectivity to better capture the complexity of pain experience.^20,21,22^ In light of this network-based view of pain, white matter hyperintensities (WMH) emerge as a potential structural marker of disrupted communication within pain-related brain networks. WMH are common in older adults and reflect vascular pathology due to cerebral small vessel disease.^63^ They have been associated with deficits in cognition, motor coordination, and emotional regulation.^23,24,25^ Although not traditionally studied in the context of pain, recent findings suggest that WMH may alter brain function by disrupting tract-level microstructure and contributing to cortical degeneration in anatomically connected regions.^26^ These changes may impair the brain’s ability to process and regulate nociceptive input, thereby potentially influencing pain sensitivity. However, to our knowledge, no previous studies have directly examined how WMH-related network disruption contributes to individual differences in pain sensitivity, and its role therefore remains unclear.

In this study, we investigate tract-specific relationships between WMH and pain sensitivity. Based on prior evidence, we consider two structural pathways through which WMH may be associated with pain perception: microstructural alterations in pain-relevant white matter tracts, and degeneration in cortical regions structurally connected to these tracts. Using multimodal MRI and behavioral data from the population-based Rotterdam Study, we examine whether these pathways are associated with inter-individual variability in pain sensitivity. In addition, we examined these associations in the Tromsø 7 cohort of the Tromsø Study as an independent cohort for replication.

## Materials and methods

### Study population

The Rotterdam Study (RS) is a population-based cohort study of 17,931 participants aged 45 years and older, living in the same suburb of Rotterdam, the Netherlands.^27,60^ The objective of the Rotterdam Study is to investigate determinants, incidence, and progression of chronic disabling disease in the elderly. The study design and rationale are described elsewhere in detail.^27^ The first cohort, Rotterdam Study I (RS-I) started in 1989 with 7,983 participants. The study was extended in 1999 with another 3,011 individuals in Rotterdam study II (RS-II). The third cohort, Rotterdam Study III (RS-III) started in 2006 adding 3,932 participants. All participants were examined in detail at baseline and follow-up visits approximately every 6 years.

This study included participants from the RS-II cohort (fifth follow-up) and RS-III cohort (third follow-up) of the Rotterdam Study. For the present analysis, 2,068 participants had available brain MRI data. Standard image quality control procedures were applied to exclude scans with major artefacts or abnormalities that could affect image processing and segmentation, leaving 1,984 participants with high-quality T1-weighted, fluid-attenuated inversion recovery (FLAIR), and diffusion tensor imaging (DTI) data. Participants without cold pressor test (CPT) measurements or required covariate data were subsequently excluded, resulting in a final analytical sample of 1,448 participants (mean age 73 ± 6 years).

The Rotterdam Study has been approved by the Erasmus MC Medical Ethics Committee (registration number MEC 02.1015) and by the Dutch Ministry of Health, Welfare and Sport (Population Screening Act WBO, license number 1071272–159521 PG). The Rotterdam Study has been entered into the Netherlands National Trial Register (NTR; www.trialregister.nl) and into the WHO International Clinical Trials Registry Platform (ICTRP; www.who.int/ictrp/network/primary/en/) and under shared catalogue number NTR6831. All participants provided written informed consent to participate in the study and to have their information obtained from treating physicians.

### Pain measurement

Cold pressor test (CPT) tolerance time was tested by asking the participants to submerge their left hand and wrist in a vat containing circulating water held at 3°C and keep it there as long as they were able, up to a maximum of 120 seconds.^14^ The research technician instructed participants to keep their hand open and relaxed. The technician explained that the water would initially feel cold and later become painful. Participants were told that if they could no longer tolerate the discomfort, they should simply remove their hand from the water and the pain would fade within seconds. Participants were also informed that the test would be stopped after two minutes, regardless of whether they continued to tolerate the cold. A constant water temperature was ensured by continuous exchange between the vat and a circulating cooler. Participants were excluded from CPT testing if they declined to perform the test, had problems comprehending the instructions, reported medical issues that in their experience affected their response to cold (e.g., cold allergy, Raynaud syndrome, loss of sensitivity or open sores on the hand to be tested. For descriptive purposes, participants were categorized as pain-tolerant if they could endure the full 120 seconds of CPT and as pain-sensitive if they withdrew the hand from the water earlier than that. In analyses where CPT was involved, CPT duration was treated as a continuous time-to-event variable.

### Collection of potential confounding variables

Participants provided information through questionnaires covering educational attainment, smoking status, current hypertension, chronic pain, and self-reported use of pain medication in the last 24 hours. Chronic pain was determined by the question “Did you have pain anywhere in your body, for at least half of the days, during the last six weeks?” (yes/no). Blood pressure was measured by research assistants using a standardized protocol.^28^ Hypertension was defined as a resting blood pressure ≥140/90 mmHg or the use of blood pressure–lowering medication. Smoking was defined as self-reported current daily smoking. Participants with missing information on education, hypertension, chronic pain, or pain medication use were excluded from all analyses.

### MRI data collection

MRI imaging was performed on a single 1.5 Tesla MRI scanner (GE Signa Excite). The MRI protocol included a T1-weighted image (T1w, repetition time 13.8 milliseconds [ms], echo time 2.80 ms, inversion time 400 ms, 96 slices of 1.6 millimeter [mm], matrix 256 × 256), a T2-weighted fluid-attenuated inversion recovery (FLAIR) sequence (repetition time 8000 ms, echo time 120 ms, inversion time 2000 ms, 64 slices of 2.5 mm, matrix 320 × 224). For diffusion weighted imaging (DWI), we performed a single shot, diffusion-weighted spin-echo echo-planar imaging sequence (repetition time (TR) = 8575 ms, echo time (TE) = 82.6 ms, axial field-of-view (FOV) = 210 × 210 mm, matrix = 96 × 64 (phase encoding) (zero-padded in k-space to 256 × 256) slice thickness = 3.5 mm, 35 contiguous slices). Maximum b-value was 1000 s/mm^2^ in 25 noncollinear directions; three volumes were acquired without diffusion weighting (b-value = 0 s/mm^2^).^29^

### MRI data preprocessing

Subcortical volume (41 regions), cortical volume (68 regions), cortical thickness (68 regions) and intracranial volume (ICV) were derived from the T1-weighted images using FreeSurfer version 6.0.0.^30^ Diffusion tensor imaging (DTI) data were converted from DICOM to NIfTI format using dcm2nii,^31^ after which they were processed using the FSL pipeline to derive fractional anisotropy (FA) and mean diffusivity (MD) maps.^32^ All FA and MD maps were nonlinearly registered to the standard-space FMRIB58_FA template (1 × 1 × 1 mm³, MNI152 space) using FNIRT.^32^ Predefined white matter tract templates from the Rotterdam Study were used,^33^ comprising 27 white matter tracts (including left and right hemispheric tracts). A 90% probability threshold was applied to define tract masks, and mean FA and MD values within each tract were then extracted for each participant. We applied the PGS pipeline,^34^ a trained deep learning-based approach, to automatically segment the WMH from the co-registered T1-weighted and FLAIR images. A white matter mask, which was derived from FreeSurfer for each individual scan, was applied to the deep learning-based WMH segmentation results, so as to remove any non-white-matter artifacts in the WMH segmentation outcomes. The T1-weighted images were nonlinearly registered to the MNI152 T1 template using FNIRT, and the resulting transformation was applied to the segmented WMH masks to bring them into MNI space. WMH volume within each of the 27 predefined tracts was then calculated.

To identify tract-connected cortical regions, each predefined tract mask in MNI space was intersected voxel-wise with the Harvard–Oxford cortical atlas,^32^ and cortical regions containing overlapping voxels were defined as tract-connected cortical regions.

### Statistical analysis

We conducted tract-specific regression analyses to examine associations between WMH burden and pain sensitivity. For the analysis of CPT, we conducted survival analysis using the Cox proportional hazards model.^35^ The event was defined as early withdrawal of the hand before the end of the test (coded as 1), while participants who reached the maximum test duration were treated as censored observations (coded as 0), with the maximum duration set to 119 seconds (instead of 120 seconds) to account for minor procedural variation in data collection; Time-to-event was the CPT duration for each participant; Independent variables were the tract-specific WMH volume, defined in MNI space; Covariates included age, sex, total WMH volume, and ICV. To assess the contribution of potential confounders, we performed sensitivity analyses including additional covariates such as chronic pain (yes/no based on questionnaire), and pain medication use (yes/no), hypertension (self-reported), educational years (continuous), and smoking (current daily smoking). Detailed definitions of these variables are provided in the section *Collection of potential confounding variables*, and the detailed results of these sensitivity analyses are provided in the Supplementary Figure S2. In addition, we performed separate regression analyses to examine (1) the association between tract-specific DTI metrics and pain sensitivity using Cox regression, and (2) the association between tract-specific WMH volume and mean FA and MD within the same tract. To further examine the proposed brain–pain pathway, we performed mediation analyses within a Cox proportional hazards framework to test whether WMH may influence pain sensitivity through DTI or cortical changes.^35,36^ We considered WMH volume within a tract as the exposure variable of the pathway, focusing on tracts that showed (nominal)significant associations in the Cox proportional hazards model. CPT duration was used as the time variable, with early withdrawal defined as the event. Covariates included age, sex, total WMH volume, and ICV. We considered two potential indirect pathways and evaluated them in separate mediation models: 1) tract-specific DTI FA as a mediator, and 2) tract-connected cortical measurements as mediators.

In all analyses, multiple testing correction was performed across the tracts using the false discovery rate (FDR) approach to control for the expected proportion of false positives among statistically significant findings.^37^ FDR-adjusted p-values (FDR-p) were calculated using the Benjamini–Hochberg procedure. Statistical significance was defined as FDR-p < 0.05, while nominal p-values are reported for completeness. For all regression analyses, tract-specific WMH volumes were first log-transformed to reduce skewness, after which tract-specific WMH, FA, and MD measures were rescaled to a 0–1 range using min–max normalization.

### Replication analysis

We conducted replication analysis using an external population-based cohort, the Tromsø 7 cohort of the Tromsø Study. The Tromsø Study is a population-based health study conducted in Tromsø, Norway since 1974. Detailed study design, recruitment procedures, MRI acquisition, pain assessment, and collection of potential confounding variables have been described previously.^14,59^ In the present study, we included 1,522 participants aged 40–84 years who underwent brain MRI, completed the cold pressor test (CPT), and had complete data for the primary analyses.^14^ Use of data from the Tromsø Study was approved by the Regional Committee for Medical and Health Research Ethics, Health Region North, reference number 216989.

The MRI data preprocessing followed the same settings as the discovery cohort (Rotterdam Study) described in Method Section *MRI data preprocessing*. However, DTI data, which were available in the Rotterdam Study, were absent in the Tromsø Study. To enable tract-level WMH analysis, predefined white matter tract templates comprising 27 tracts, derived from Rotterdam Study DTI data and defined in MNI space (MNI152),^33^ were applied. WMH were segmented in Tromsø Study and subsequently registered to the MNI space. In this way, the Rotterdam-derived tract templates were applied to the Tromsø WMH maps to quantify tract-specific WMH volumes.

For statistical analysis, we replicated the Cox-regression analysis between tract-specific WMH volumes and CTP. We also replicated the mediation analyses, for the indirect pathway considering tract-connected cortical measurements as potential mediators. Effect estimates from the discovery cohort (Rotterdam Study) and the replication cohort (Tromsø Study) were combined using an inverse-variance weighted fixed-effect meta-analysis based on summary statistics.

## Results

### Population characteristics

Characteristics of the study population of the Rotterdam Study are shown in Table 1. In total, 1,448 participants were included in the analysis. The mean age was 73 ± 6 years, and 53% were females. In the cold pressor test (CPT), 411 (28%) participants reached the maximum test duration of 120 seconds, while 1,037 (72%) participants withdrew their hand earlier (Figure 2). Among those who withdrew early, the mean CPT duration was 33.9 ± 21.8 seconds. Compared with pain-tolerant participants, the pain-sensitive group included a higher proportion of females and greater use of pain medication within 24 hours prior to the CPT. Educational level also differed between the groups, with lower educational attainment in the pain-sensitive group, whereas age, smoking status, chronic pain prevalence, and hypertension were comparable.

**Figure 1:**
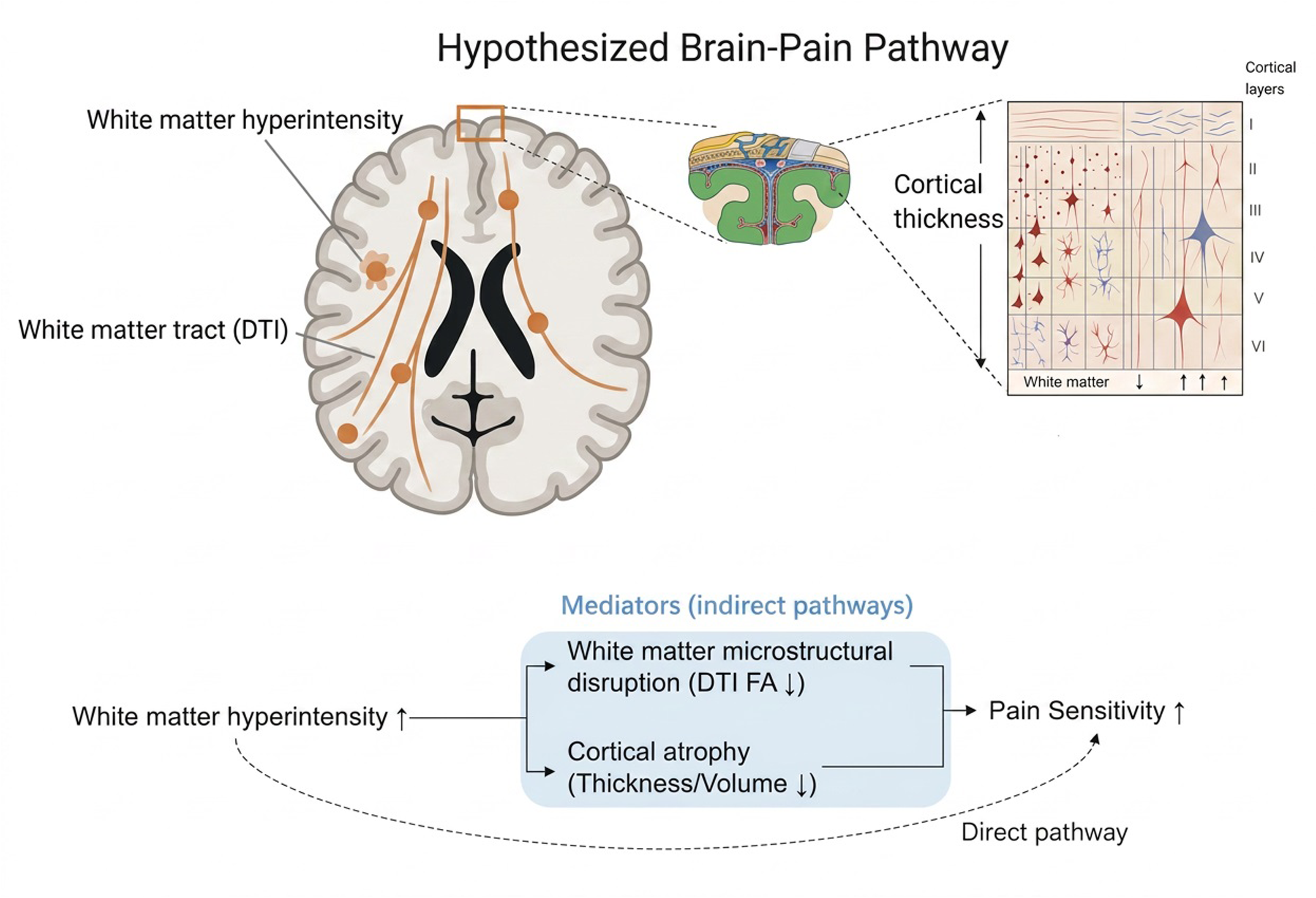
Hypothesized brain-pain pathway. White matter hyperintensities (WMH) may increase pain sensitivity through both direct and indirect pathways. Indirect effects are mediated by (1) disruption of white matter microstructure, reflected by reduced diffusion tensor imaging (DTI) fractional anisotropy (FA), and (2) cortical atrophy in regions structurally connected to affected tracts. These pathways represent complementary mechanisms linking vascular white matter damage to altered pain processing.

**Figure 2:**
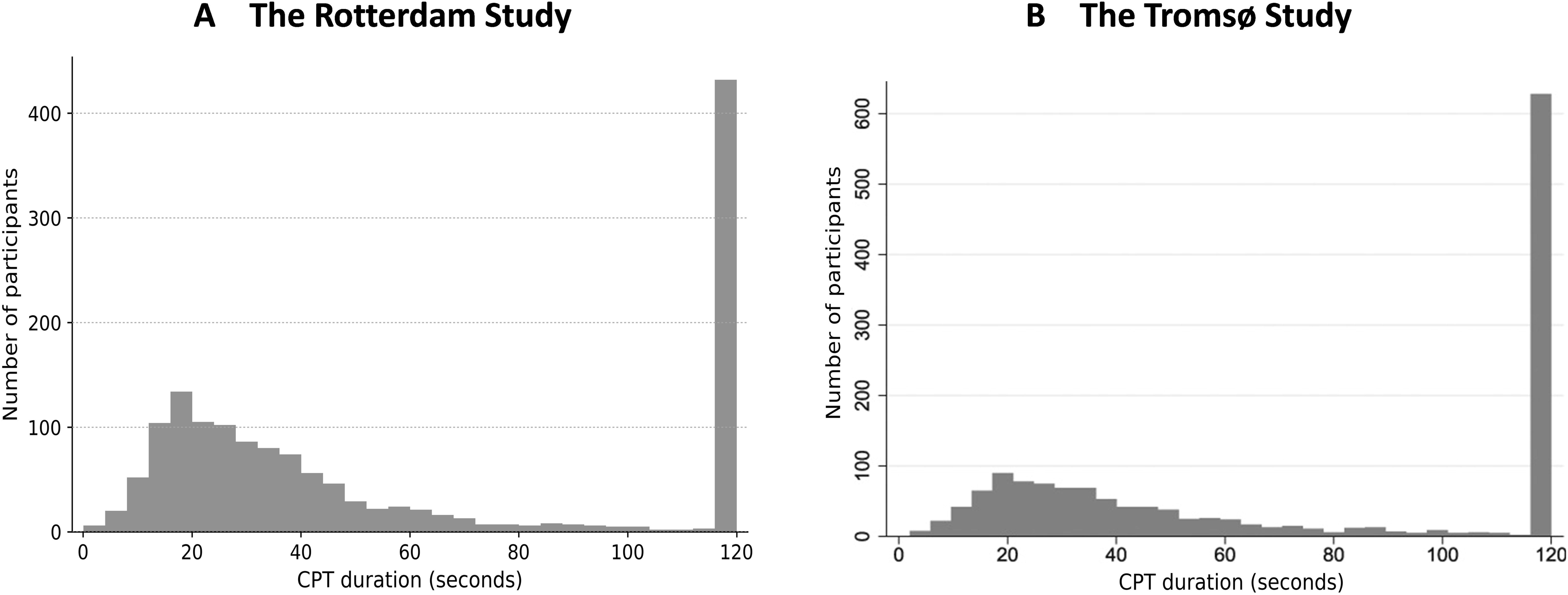
Histogram of cold pressor test (CPT) duration of the participants. **(A)** The Rotterdam Study; **(B)** The Tromsø Study.

**Table 1:**
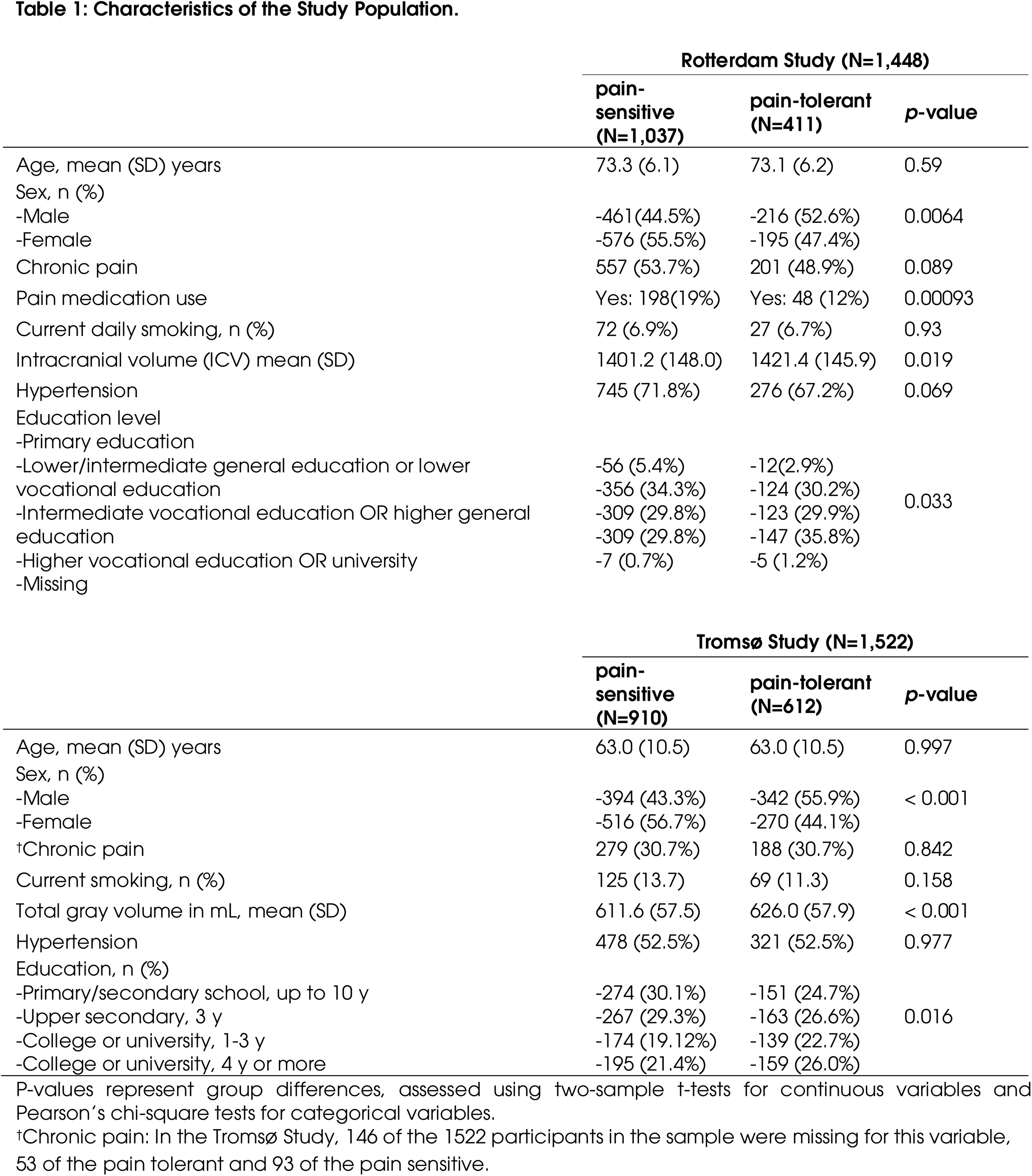
Characteristics of the Study Population.

The characteristics of the replication cohort from the Tromsø Study are summarized in Table 1 and Figure 2. In total, 1,522 participants were included in the analysis. Among them, 612 (40.2%) were classified as pain tolerant, reaching the maximum cold pressor test (CPT) duration of 120 seconds, whereas 910 (59.8%) withdrew their hand earlier and were classified as pain sensitive. The mean age of participants was 63.0 ± 10.5 years. Compared with pain-tolerant participants, the pain-sensitive group included a higher proportion of females and slightly lower total gray matter volume, while education level also differed between the two groups. Chronic pain prevalence and hypertension were comparable between groups.

### Global-level analysis between brain measurement and CPT, in the Rotterdam Study

At the global level, smaller total brain, gray matter, and white matter volumes were linked with greater pain sensitivity, although these associations did not reach statistical significance (Supplementary Table S1). In contrast, greater total WMH volume showed a marginal association with higher pain sensitivity (nominal *p* = 0.061), which reached statistical significance for the left hemisphere (nominal *p* = 0.047). Mean FA demonstrated a trend toward lower values being associated with greater pain sensitivity, particularly in the left hemisphere (nominal *p* = 0.050).

### Tract-specific WMH volume associated with tract-specific DTI metrics, in the Rotterdam Study

White matter hyperintensities (WMH) were detected in 20 of the 27 predefined tracts, except for the left and right cingulum (hippocampal part; CGH), left and right fornix (FX), left and right medial lemniscus (ML), and the middle cerebellar peduncle (MCP). Figure 3a shows the distribution of WMH across the 27 predefined tracts, with each bar representing the WMH percentage relative to the tract’s total volume (100% indicating complete tract coverage. Highest relative WMH burden was seen in the forceps major tract (FMA, 2.05%), left and right anterior thalamic radiation (ATR_L, 1.86%), and ATR_R (1.51%). Overall, WMH percentages were slightly higher in the left tracts compared to their right homologues.

**Figure 3:**
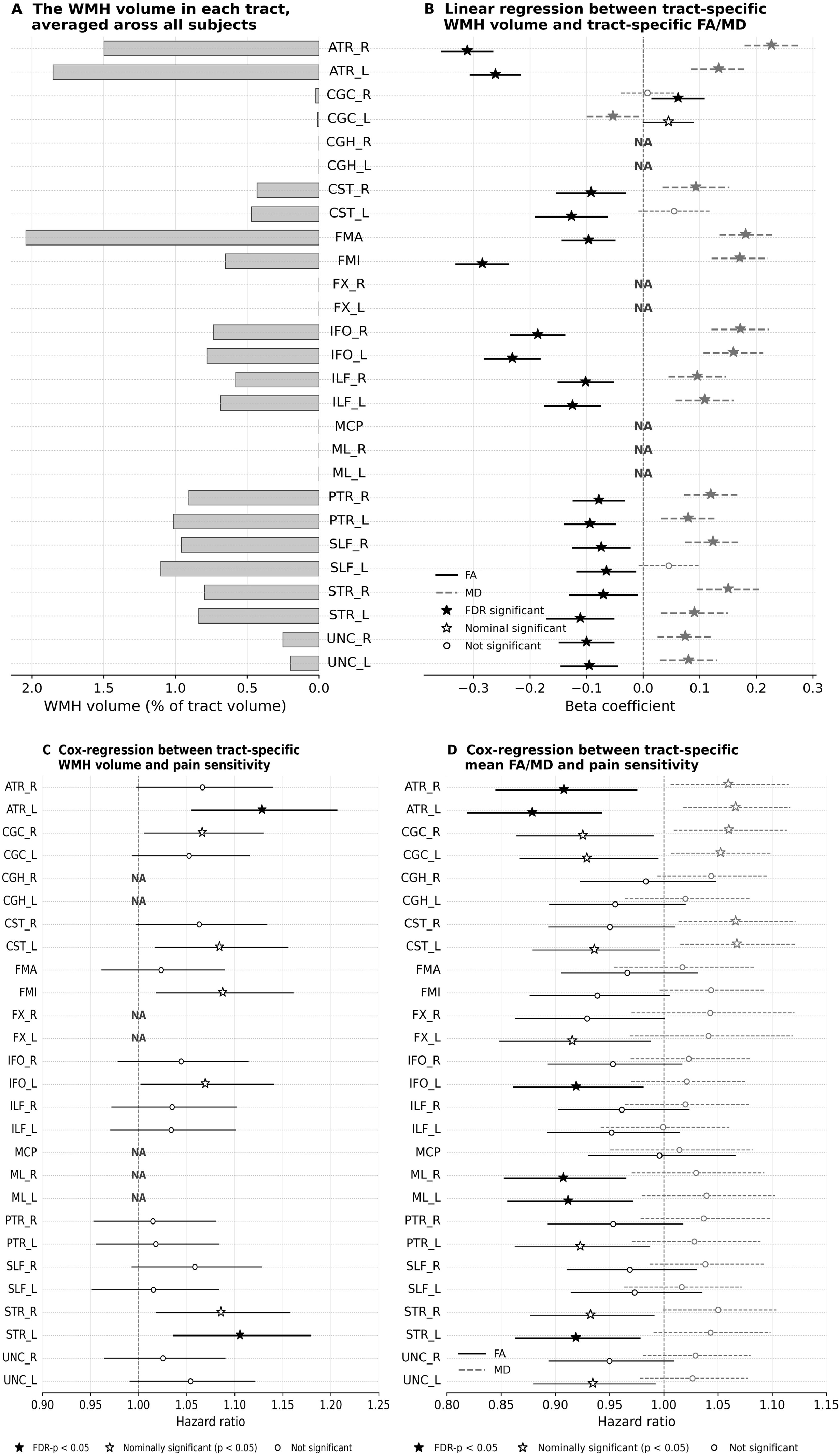
Tract-specific associations between WMH, white matter microstructure, and pain sensitivity in the Rotterdam Study. **(a)** Bar chart showing the distribution of white matter hyperintensities (WMH) burden across 27 predefined tracts, with each bar representing the WMH percentage relative to the tract’s total volume (100% indicating complete tract coverage); **(b)** Association between tract-specific WMH and tract-specific fractional anisotropy (FA) and mean diffusivity (MD), based on linear regression analysis, adjusted for age, sex, total WMH volume, and intracranial volume (ICV); **(c)** Cox-regression analysis between cold pressor test (CPT) performance and tract-specific white matter hyperintensity (WMH), adjusted for age, sex, total WMH volume, and ICV; **(d)** Cox-regression analysis between CPT performance and tract-specific fractional anisotropy (FA) and mean diffusivity (MD), adjusted for age, sex, and ICV. For panels (**b), (c), and (d**), WMH was log-transformed. WMH, FA, and MD measures were rescaled to a 0–1 range using min–max normalization prior to regression analyses. Abbreviations: ATR, anterior thalamic radiation; IFO, inferior fronto-occipital fasciculus; ILF, inferior longitudinal fasciculus; PTR, posterior thalamic radiation; SLF, superior longitudinal fasciculus; UNC, uncinate fasciculus; FMA, forceps major; FMI, forceps minor; CGC, cingulate gyrus part of cingulum; CGH, parahippocampal part of cingulum; CST, corticospinal tract; MCP, middle cerebellar peduncle; ML, medial lemniscus; STR, superior thalamic radiation; FX, fornix. _L and _R denote left and right hemispheres, respectively. NA denotes tracts with no WMH detected and therefore no analysis performed. FDR: false discovery rate. The Rotterdam Study.

Regression analysis indicated that higher WMH burden within 18 out of the 20 WMH-detected tracts was significantly associated with reduced fractional anisotropy (FA) within the same tract (Figure 3b). Inconsistent patterns were observed in the left and right cingulate gyrus part of cingulum (CGC_L, CGC_R) tracts, in which higher WMH volume was associated with higher FA. These two tracts had relatively low WMH burden at 0.01% and 0.02%, respectively. Complementary analyses of mean diffusivity (MD) showed associations in the opposite direction, with higher WMH burden associated with increased MD across the same tracts, except for the left and right inferior longitudinal fasciculus (ILF_L, ILF_R), which were not statistically significant.

### Tract-specific WMH and DTI metrics associated with pain sensitivity, in the Rotterdam Study

In the association analysis with pain sensitivity, Cox regression was performed using CPT duration as the time variable, with early withdrawal defined as the event. Lower FA, which was associated with higher WMH burden (Figure 3b), was also associated with greater pain sensitivity (Figure 3d), particularly in the left and right anterior thalamic radiation tract (ATR_L: FDR-*p* < 0.0081; ATR_R: FDR-*p* <0.042), the left and right medial lemniscus tract (ML_L: FDR-*p* <0.035; ML_R: FDR-*p* <0.026), the left superior thalamic radiation tract (STR_L, FDR-*p* < 0.042), and the left inferior fronto-occipital fasciculus tract (IFO_L: FDR-*p* <0.048). MD showed nominal associations in the opposite direction, with higher MD associated with greater pain sensitivity, but these did not survive multiple testing correction.

In addition, higher WMH burden within a tract was directly associated with higher pain sensitivity (Figure 3c), particularly in the ATR_L tract (FDR-*p* <0.0077), and left superior thalamic radiation tract (STR_L, FDR-*p* < 0.023). “NA” in Figure 3 denotes tracts with no detectable WMH and therefore no analysis performed.

### Mediation analysis in the Rotterdam Study

Figure 4 maps the mediation analysis results on a brain atlas, with DTI metrics (Figure 4a) and cortical measures (Figure 4b) evaluated as mediators in separate analyses, highlighting brain tracts and regions identified in the mediation analysis; corresponding results are summarized in Table 2 (DTI mediators) and Table 3 (cortical mediators).

**Figure 4.**
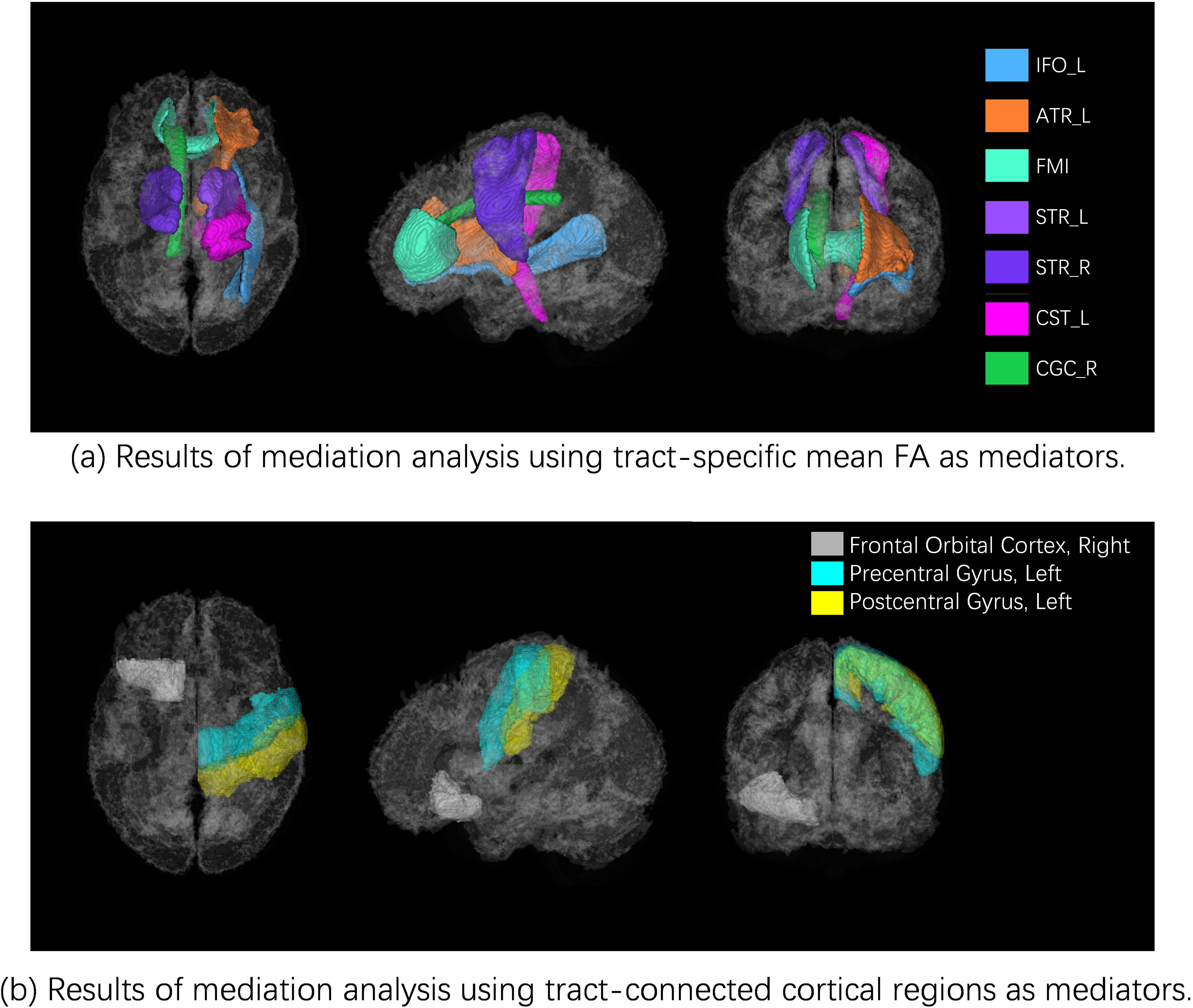
Brain atlas visualization of nominal significant (p<0.05) findings in mediation analysis in the Rotterdam Study. Pars orbitalis was approximated by the frontal orbital cortex in the Harvard–Oxford atlas.^32^ Abbreviations: IFO: Inferior fronto-occipital fasciculus; ATR: Anterior thalamic radiation; FMI, forceps minor; STR: Superior thalamic radiation; CST: Corticospinal tract; CGC, cingulate gyrus part of cingulum. _L and _R denote left and right hemispheres, respectively. The Rotterdam Study.

**Table 2:**
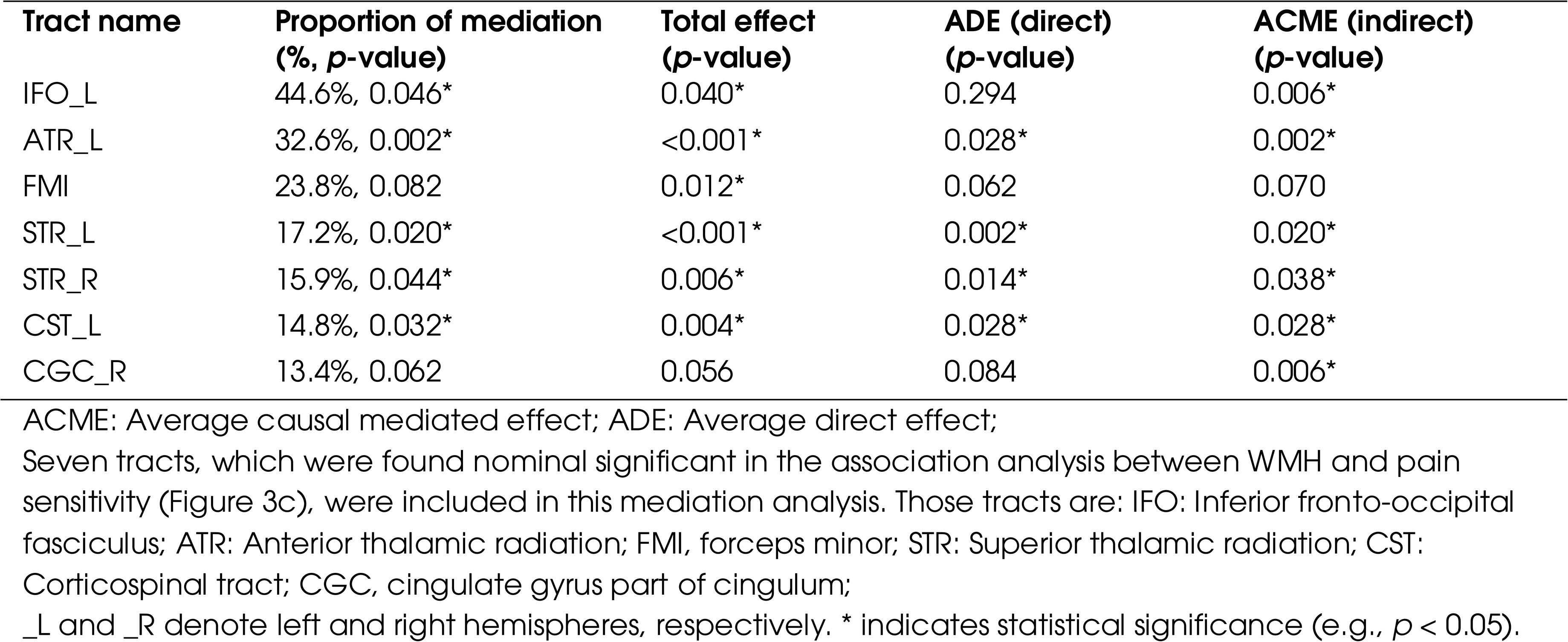
Mediation analysis using tract-specific fractional anisotropy as the potential mediator, in the Rotterdam Study. Exposure variable: log-transformed WMH volume within a tract; Outcome variable: CPT, analysed as a time-to-event measure of pain sensitivity. The Rotterdam Study.

| Tract name | Proportion of mediation<br>(%, *p*-value) | Total effect<br>(*p*-value) | ADE (direct)<br>(*p*-value) | ACME (indirect)<br>(*p*-value) |
| --- | --- | --- | --- | --- |
| IFO_L | 44.6%, 0.046* | 0.040* | 0.294 | 0.006* |
| ATR_L | 32.6%, 0.002* | <0.001* | 0.028* | 0.002* |
| FMI | 23.8%, 0.082 | 0.012* | 0.062 | 0.070 |
| STR_L | 17.2%, 0.020* | <0.001* | 0.002* | 0.020* |
| STR_R | 15.9%, 0.044* | 0.006* | 0.014* | 0.038* |
| CST_L | 14.8%, 0.032* | 0.004* | 0.028* | 0.028* |
| CGC_R | 13.4%, 0.062 | 0.056 | 0.084 | 0.006* |

ACME: Average causal mediated effect; ADE: Average direct effect;
\_L and \_R denote left and right hemispheres, respectively. \* indicates statistical significance (e.g., $p < 0.05$ ).

**Table 3:**
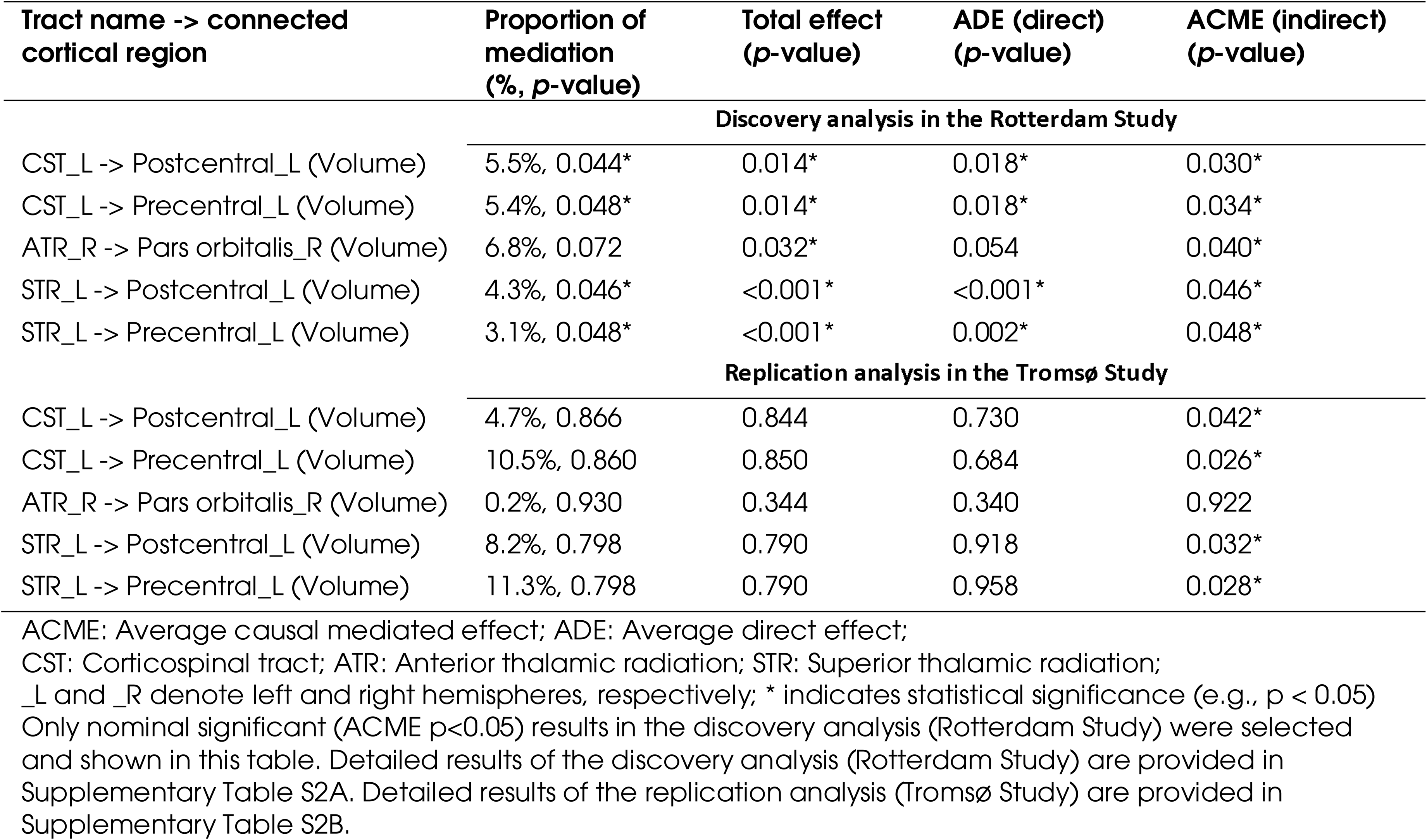
Mediation analysis using tract-connected cortical regions as potential mediators. Exposure variable: log-transformed WMH volume within a tract; Outcome variable: CPT, analysed as a time-to-event measure of pain sensitivity.

In Table 2, mediation analysis indicated that the association between tract-specific WMH burden and pain sensitivity was partially statistically mediated by reduced FA within several white matter tracts. Among the seven tracts showing nominal associations (*p* < 0.05) in the primary Cox regression (Figure 3c), largest relative proportion mediated was observed in the left inferior fronto-occipital fasciculus (IFO_L; 44.6% mediation, average causal mediated effect [ACME] *p* = 0.006) and the left anterior thalamic radiation (ATR_L; 32.6% mediation, ACME *p* < 0.002). More modest but statistically significant mediation effects were detected in the left superior thalamic radiation (STR_L; 17.2%, ACME *p* = 0.020), right superior thalamic radiation (STR_R; 15.9%, ACME *p* = 0.038), left corticospinal tract (CST_L; 14.8%, ACME *p* = 0.028), and right cingulum (CGC_R; 13.4%, ACME *p* = 0.006). For the forceps minor (FMI), the mediation proportion was 23.8%, but the indirect effect did not reach statistical significance (ACME *p* = 0.070).

Table 3 shows that cortical atrophy demonstrated significant indirect effects in the mediation analysis, accounting for a smaller proportion of the total mediation effect. In particular, the association between WMH burden in the left corticospinal tract (CST_L) and greater pain sensitivity was partially mediated by cortical atrophy in two cortical regions. Degeneration in the left postcentral gyrus mediated 5.3% of the total effect (ACME *p* = 0.048), while degeneration in the left precentral gyrus mediated 5.9% of the total effect (ACME *p* = 0.052). In both pathways, the direct effects were also statistically significant (Average Direct Effect [ADE] *p* = 0.018 and p = 0.014, respectively). Additional indirect effects were observed for the superior thalamic radiation (STR_L), with cortical atrophy in also the left postcentral and left precentral regions mediating 4.1% (ACME *p* = 0.034) and 3.0% (ACME *p* = 0.076) of the association. Complete results for other tracts and cortical regions are provided in Supplementary Table S2A.

### Replication analysis in the Tromsø Study

Supplementary Figure S1 shows the distribution of WMH across the 27 predefined tracts in the Tromsø Study, demonstrating a pattern consistent with the Rotterdam Study, with the highest WMH burden in FMA (2.17%) and ATR_L (1.59%).

Figure 5 shows the replication results from the Tromsø Study for the association between CPT performance and tract-specific WMH volume based on Cox regression analysis. In the Tromsø Study, higher WMH burden within a tract was generally associated with higher pain sensitivity, particularly in the forceps major (FMI; nominal *p* < 0.024), consistent with the findings from the Rotterdam Study (FMI; nominal *p* < 0.012). After meta-analysis, the increased statistical power led to four tracts reaching statistical significance after multiple testing correction: ATR_L (FDR-*p* < 0.006), CGC_R (FDR-*p* < 0.033), FMI (FDR-*p* < 0.006), and STR_L (FDR-*p* < 0.030).

**Figure 5:**
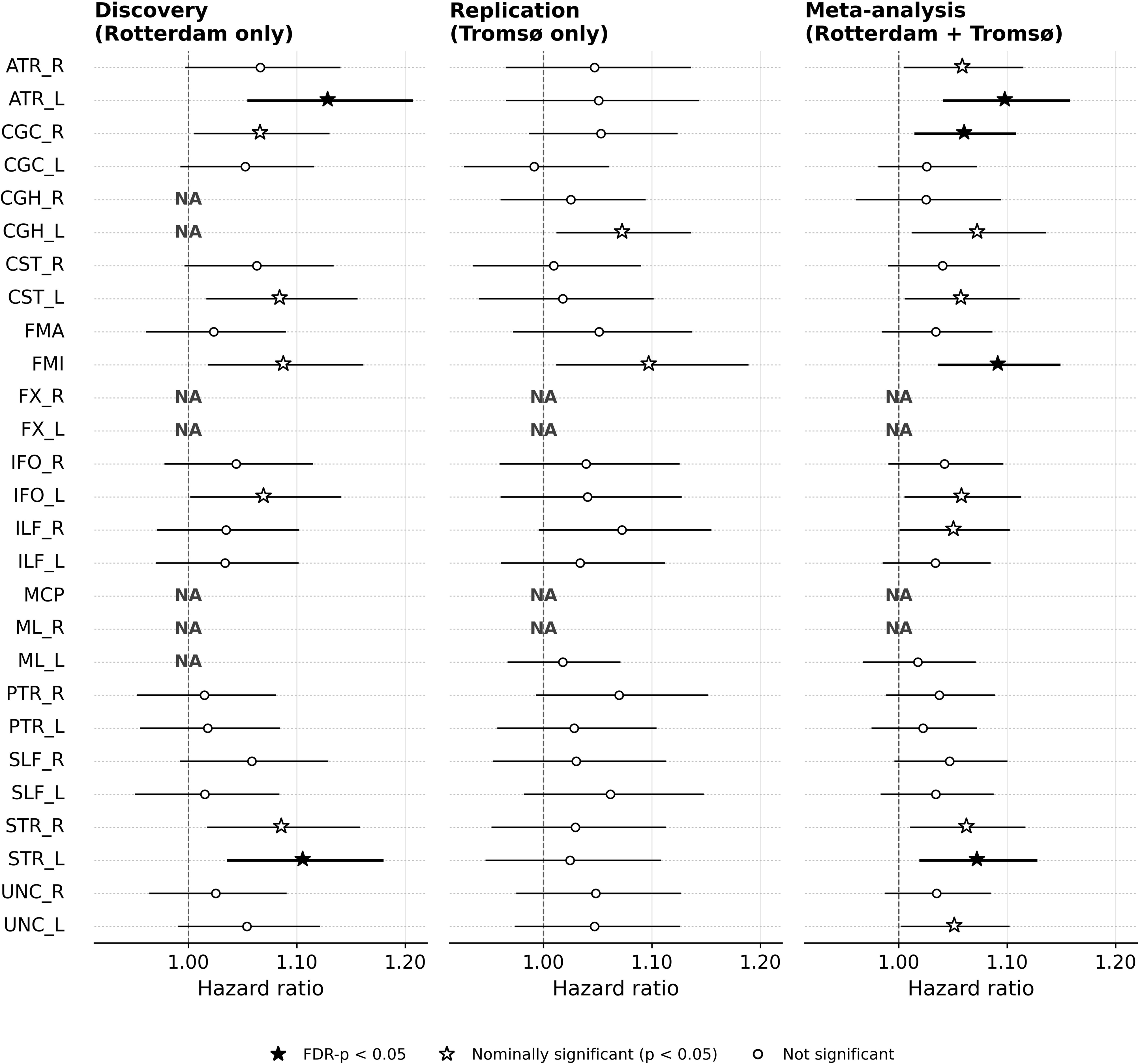
Replication and meta-analysis of tract-specific WMH associations with pain sensitivity. Replication results from the Tromsø Study for the association between cold pressor test (CPT) performance and tract-specific white matter hyperintensity (WMH) volume based on Cox regression analysis, and meta-analysis results combining Rotterdam Study and Tromsø Study. Covariates included age, sex, total WMH volume, and intracranial volume (ICV). WMH was log-transformed and normalized to a 0–1 scale. Abbreviations: ATR, anterior thalamic radiation; IFO, inferior fronto-occipital fasciculus; ILF, inferior longitudinal fasciculus; PTR, posterior thalamic radiation; SLF, superior longitudinal fasciculus; UNC, uncinate fasciculus; FMA, forceps major; FMI, forceps minor; CGC, cingulate gyrus part of cingulum; CGH, parahippocampal part of cingulum; CST, corticospinal tract; MCP, middle cerebe llar peduncle; ML, medial lemniscus; STR, superior thalamic radiation; FX, fornix. _L and _R denote left and right hemispheres, respectively. FDR: false discovery rate.

Table 3 presents the replication results of the mediation analysis examining tract-connected cortical measurements as potential mediators of the association between tract-specific WMH volume and CPT performance. Although the total effect was not significant in the Tromsø Study, significant indirect effects (ACME *p* < 0.05) were observed in the left precentral and left postcentral cortical regions. These two regions mediated the association for both the corticospinal tract (CST) and superior thalamic radiation (STR), consistent with the pathways identified in the discovery cohort (Table 3).

## Discussion

This study is, to our knowledge, the first to map tract-specific associations between white matter hyperintensities (WMH) and pain sensitivity. After multiple-testing correction, higher WMH burden and lower fractional anisotropy (FA) were each associated with greater pain sensitivity across multiple tracts. Mediation analyses suggested that the relationship between WMH burden and pain sensitivity was primarily explained by tract-level microstructural alterations, particularly within the anterior thalamic radiations (ATR). In contrast, cortical atrophy showed comparatively modest indirect effects, mainly involving the precentral and postcentral gyri, which also demonstrated nominally significant mediation effects in the replication cohort. Together, these findings support the notion that WMH may be linked to increased pain sensitivity through structural alterations in white matter tracts, with a smaller additional contribution from cortical atrophy in connected regions.

When comparing the two identified pathways, the stronger mediation effect observed for DTI-based microstructural changes compared with cortical atrophy likely reflects both the temporal dynamics and the anatomical spread of WMH-related damage. Diffusion metrics such as FA are highly sensitive to subtle microstructural alterations that occur in parallel with, or even before, visible WMH formation. Longitudinal work in aging and small vessel disease has shown that lower baseline FA in normal-appearing white matter predicts later WMH development, supporting the view that tract integrity deteriorates early and directly in response to vascular pathology.^38,39^ Similarly, studies combining tractography and cortical morphometry suggest that WMH are associated with local FA reductions along connected tracts, and that disruption of white matter pathways may contribute to cortical thinning in anatomically connected regions.^40,41^ Consistent with this, lesion-based longitudinal studies further demonstrate secondary cortical thinning in remote, tract-connected regions following focal white matter injury.^42^ This may reflect that DTI captures more proximal tract-level alterations, whereas cortical thinning reflects more downstream and distributed consequences of disrupted connectivity. Such a mechanistic pattern explains the large and robust mediation effects observed for the DTI pathway (e.g., 32.6% in the left anterior thalamic radiation), in contrast to the weaker effects observed for cortical mediation (all <6.8%), where only a subset of tract–region pairs reached statistical significance. An additional explanation relates to anatomical divergence. Given that individual white matter tracts project to multiple cortical regions, WMH-related disruption may be distributed across several connected regions rather than concentrated in a single cortical area. Consequently, mediation estimates at the level of individual cortical regions may be attenuated or less consistent. Taken together with prior literature, these results suggest a biologically plausible model in which WMH primarily disrupts white matter microstructure, with cortical atrophy reflecting a more delayed and spatially dispersed downstream consequence. Although our design is cross-sectional, our findings are consistent with longitudinal studies demonstrating that microstructural alterations in normal-appearing white matter precede WMH development and expansion,^38,39^ and with evidence linking WMH-related disconnection to subsequent cortical atrophy.^40,41,42^

The identified pathways reflect both mental and physical components of pain processing. The tract-specific mediation analyses identified several key tract-specific pathways, including the thalamic radiations (anterior, superior, and posterior; ATR, STR, PTR) and the corticospinal tract (CST). The medial lemniscus (ML) showed a direct association between lower FA and increased pain sensitivity, but no WMH burden was present in this tract to enable mediation testing. Together, these tracts represent complementary pathways that are involved in distinct yet interrelated aspects of pain processing. The thalamic radiations form the principal communication routes between the thalamus and the cortex, supporting the integration of cognitive, emotional, and sensory information crucial for pain perception.^43,61,62^ Consistent with these functions, previous studies have reported lower FA in the ATR in individuals with chronic pain conditions such as interstitial cystitis/bladder pain syndrome.^16^ In contrast, the CST and ML are central to somatosensory and motor pathways, linking sensory input with physical and behavioral pain responses. The postcentral gyrus, which is connected to both CST and ML tracts and identified in our mediation analysis, is known to be directly involved in processing sensation of touch, temperature, and pain.^44^ It is robustly activated across experimental and clinical pain paradigms, including affective evaluation of cold pain,^45^ and chronic pain conditions such as burning mouth syndrome and back pain.^46,47^ Together, these tract-specific pathways suggest that WMH may influence pain processing through both affective–cognitive (“mental”) and sensory–somatic (“physical”) components. WMH-related disruption may impair inhibitory control within pain-processing networks, leading to a relative disinhibition of pain responses. In this context, disruptions in thalamic radiations may affect top-down modulation of pain, whereas alterations in CST and ML may reflect changes in sensory processing and integration. These mechanisms may contribute to individual differences in pain tolerance and withdrawal responses during the CPT, although the precise pathways underlying these effects remain to be clarified.

The CPT, which measures tolerance to prolonged exposure to cold water, provides a multifaceted probe of pain perception. It elicits not only the sensory-discriminative aspects of cold and tactile processing but also the affective and motivational dimensions that influence an individual’s willingness to tolerate pain. This multidimensional nature may explain why our analysis identified both thalamo-cortical (mental) and sensory–motor (physical and motivational) pathways as significant correlates of pain sensitivity.

Instead of the CPT, various experimental paradigms using a range of pain assessments have been used to capture different aspects of pain perception and link them with brain MRI measurements. Thermal pain intensity was tested in 116 healthy adults by Emerson et al. 2014,^48^ showing that higher pain sensitivity was associated with lower gray matter density. Grant et al. 2010 found that Zen meditators with long-term practice exhibited lower pain sensitivity and thicker cortex,^49^ while Villemure et al. 2014 reported that yoga practitioners had greater pain tolerance and increased gray- and white-matter volumes.^50^ Similarly, Kramer et al. 2016 showed that larger insular gray-matter volume strengthened the coupling between cortical responses to heat and lower subjective pain ratings, suggesting that preserved structure facilitates more efficient pain modulation.^51^ Together, these studies, which employed supra-threshold pain stimuli with outcomes including pain ratings and pain tolerance, demonstrate that reduced brain structural integrity is associated with heightened pain sensitivity, consistent with our findings. In contrast, studies employing threshold-based measures of pain sensitivity, defined as the lowest stimulus intensity that elicits a pain sensation, have reported mixed or inconsistent associations between brain structure and pain perception. Ruscheweyh et al. 2018 tested pressure pain threshold in a population sample and found no significant associations with brain morphology.^52^ Two studies (de Kruijf et al. 2016, Neumann et al. 2021) using heat pain thresholds reported mixed directions of association across different regions.^53,54^ Schwedt and Chong 2014 used the same heat threshold paradigm and found that cortical thickness correlated negatively with pain threshold in healthy adults but positively in migraineurs,^55^ indicating reversed associations between acute and chronic pain states. These inconsistencies likely reflect differences in how pain is measured. Threshold-based tests capture the initial detection of pain, engaging different neural mechanisms from suprathreshold or tolerance measures. In contrast, tolerance-based paradigms such as the CPT may better integrate sensory and affective components, providing a more holistic indicator of overall pain sensitivity and yielding more consistent associations with brain structure.^14^

The predominantly left-hemispheric pattern observed in this study warrants careful interpretation. Notably, an inconsistency is observed when considering stimulation side. Prior studies applying stimulation to the right hand reported predominantly left-lateralized effects (Niddam et al., 2021; Schlereth et al., 2003; Jin et al., 2020),^56,57,58^ whereas in the present study stimulation was applied to the left hand, yet a similar left-lateralized pattern was observed. This inconsistency suggests that the observed hemispheric asymmetry cannot be explained solely by simple contralateral activation. One possible explanation is that pain processing may exhibit asymmetric organization that is not strictly determined by the side of stimulation, although this remains to be confirmed. While hemispheric asymmetry has been reported in previous pain imaging studies, the underlying mechanisms remain incompletely understood and may reflect more complex functional or structural organization of pain processing. Further investigation is needed to clarify the origin and significance of this lateralization.

This study presents several methodological strengths. First, it is based on a large, population-derived cohort, offering substantially greater statistical power and generalizability than most previous neuroimaging studies on pain sensitivity. Second, pain was measured using the cold pressor test (CPT), a well-validated and reproducible tolerance paradigm that captures both sensory and affective dimensions of pain perception. Third, the integration of multimodal MRI data, including FLAIR, diffusion tensor imaging, and cortical morphometry, together with a tract-specific analytical framework, enabled a comprehensive, tract-level assessment of structural pathways linking WMH, tract integrity, and cortical morphology. Importantly, key findings were further evaluated and confirmed in an independent population-based cohort (the Tromsø Study), strengthening the robustness and reproducibility of the observed associations. This mechanistic pattern is consistent with prior evidence in vascular and neurodegenerative conditions, underscoring the broader applicability and biological relevance of the identified pathways.

This study has several limitations. Its cross-sectional design precludes causal inference, and longitudinal data are needed to confirm the temporal sequence of WMH-related pathways. White matter tracts were defined using a predefined atlas rather than subject-specific tractography, which may limit the ability to capture individual variability, particularly in the Tromsø cohort where DTI data were not available. The older age of the study population may also limit generalizability to younger or clinical cohorts with different pain profiles. In addition, the discovery and replication cohorts differed in demographic characteristics, particularly age and geographic setting, which may introduce heterogeneity when comparing results across the two populations. Such differences may influence pain perception and tolerance, as environmental and cultural factors, including habitual exposure to colder climates, could affect responses to the CPT and thereby impact the comparability of results across cohorts.

In conclusion, these findings indicate that pain sensitivity in older adults is closely linked to the structural integrity of brain circuits involved in sensory, cognitive, and emotional processing. Vascular-related white matter damage appears to play a central role in shaping how pain is encoded and tolerated at the level of the central nervous system. This provides a potential explanation for the heightened pain sensitivity observed in individuals with greater white matter hyperintensity burden, even in the absence of marked peripheral disease. Our findings extend current understanding of the neurobiological basis of pain sensitivity by linking tract-specific white matter pathology to pain processing and identifying mediation pathways. They highlight the importance of central brain mechanisms in pain assessment and may inform future research and clinical strategies.

## Data availability

Data availability is restricted due to their sensitive nature. Deidentified data can be obtained by application to the Tromsø study. Contact for details.

Access to individual-level data from the Rotterdam Study cohorts used in this study is restricted to researchers in direct collaboration with the cohort owners. Collaboration requests can be directed to Frank J.A. van Rooij at Erasmus MC;

## Acknowledgements

The Rotterdam Study is funded by the Erasmus Medical Center and Erasmus University, Rotterdam, Netherlands Organization for the Health Research and Development (ZonMw), the Research Institute for Diseases in the Elderly, the Ministry of Education, Culture and Science, the Ministry for Health, Welfare and Sports, the European Commission (Directorate-General XII), and the Municipality of Rotterdam. The authors are grateful to the study participants, the staff from the Rotterdam Study, and the participating general practitioners and pharmacists.

This work has also been submitted as an abstract and presented as an oral presentation at the ESMRMB conference in Marseille, France, on October 10, 2025.

## Supplementary material

Supplementary material is available at *Brain* online.

## References

1. Nielsen CS, Staud R, Price DD. Individual differences in pain sensitivity: measurement, causation, and consequences. J Pain. 2009 Mar;10(3):231–7.

2. Øhrn AM, Nielsen CS, Schirmer H, Stubhaug A, Wilsgaard T, Lindekleiv H. Pain Tolerance in Persons With Recognized and Unrecognized Myocardial Infarction: A Population-Based, Cross-Sectional Study. J Am Heart Assoc. 2016 Dec 21;5(12):e003846.

3. Stabell N, Stubhaug A, Flægstad T, Nielsen CS. Increased pain sensitivity among adults reporting irritable bowel syndrome symptoms in a large population-based study. Pain. 2013 Mar;154(3):385–392.

4. Staud R, Weyl EE, Price DD, Robinson ME. Mechanical and heat hyperalgesia highly predict clinical pain intensity in patients with chronic musculoskeletal pain syndromes. J Pain. 2012 Aug;13(8):725–35

5. Suokas AK, Walsh DA, McWilliams DF, Condon L, Moreton B, Wylde V, Arendt-Nielsen L, Zhang W. Quantitative sensory testing in painful osteoarthritis: a systematic review and meta-analysis. Osteoarthritis Cartilage. 2012 Oct;20(10):1075–85

6. Samuelsen PJ, Nielsen CS, Wilsgaard T, Stubhaug A, Svendsen K, Eggen AE. Pain sensitivity and analgesic use among 10,486 adults: the Tromsø study. BMC Pharmacol Toxicol. 2017 Jun 9;18(1):45.

7. Kasch H, Qerama E, Bach FW, Jensen TS. Reduced cold pressor pain tolerance in non-recovered whiplash patients: a 1-year prospective study. Eur J Pain. 2005 Oct;9(5):561–9.

8. Apkarian AV, Bushnell MC, Treede RD, Zubieta JK. Human brain mechanisms of pain perception and regulation in health and disease. Eur J Pain. 2005 Aug;9(4):463–84.

9. Coghill RC. The Distributed Nociceptive System: A Framework for Understanding Pain. Trends Neurosci. 2020 Oct;43(10):780–794.

10. Duerden EG, Albanese MC. Localization of pain-related brain activation: a meta-analysis of neuroimaging data. Hum Brain Mapp. 2013 Jan;34(1):109–49

11. Tracey I, Mantyh PW. The cerebral signature for pain perception and its modulation. Neuron. 2007 Aug 2;55(3):377–91.

12. Duerden EG, Albanese MC. Localization of pain-related brain activation: a meta-analysis of neuroimaging data. Hum Brain Mapp 2013;34:109–49.

13. Apkarian AV, Bushnell MC, Treede RD, Zubieta JK. Human brain mechanisms of pain perception and regulation in health and disease. Eur J Pain 2005;9:463–84.

14. Melum TA, Vangberg TR, Johnsen LH, Steingrímsdóttir ÓA, Stubhaug A, Mathiesen EB, Nielsen C. Gray matter volume and pain tolerance in a general population: the Tromsø study. Pain. 2023 Aug 1;164(8):1750–1758.

15. McDonagh MS, Selph SS, Buckley DI, Holmes RS, Mauer K, Ramirez S, Hsu FC, Dana T, Fu R, Chou R. Nonopioid Pharmacologic Treatments for Chronic Pain [Internet]. Rockville (MD): Agency for Healthcare Research and Quality (US); 2020 Apr. Report No.: 20-EHC010.

16. Farmer MA, Huang L, Martucci K, Yang CC, Maravilla KR, Harris RE, Clauw DJ, Mackey S, Ellingson BM, Mayer EA, Schaeffer AJ, Apkarian AV; MAPP Research Network. Brain White Matter Abnormalities in Female Interstitial Cystitis/Bladder Pain Syndrome: A MAPP Network Neuroimaging Study. J Urol. 2015 Jul;194(1):118–26.

17. Apkarian AV, Bushnell MC, Treede RD, Zubieta JK. Human brain mechanisms of pain perception and regulation in health and disease. Eur J Pain. 2005 Aug;9(4):463–84.

18. Duerden EG, Albanese MC. Localization of pain-related brain activation: a meta-analysis of neuroimaging data. Hum Brain Mapp. 2013 Jan;34(1):109–49

19. Tracey I, Mantyh PW. The cerebral signature for pain perception and its modulation. Neuron. 2007 Aug 2;55(3):377–91

20. Kucyi A, Davis KD. The dynamic pain connectome. Trends Neurosci. 2015 Feb;38(2):86–95.

21. Neumann L, Wulms N, Witte V, Spisak T, Zunhammer M, Bingel U, Schmidt-Wilcke T. Network properties and regional brain morphology of the insular cortex correlate with individual pain thresholds. Hum Brain Mapp. 2021 Oct 15;42(15):4896–4908.

22. Spisak T, Kincses B, Schlitt F, Zunhammer M, Schmidt-Wilcke T, Kincses ZT, Bingel U. Pain-free resting-state functional brain connectivity predicts individual pain sensitivity. Nat Commun. 2020 Jan 10;11(1):187.

23. Sachdev PS, Wen W, Christensen H, Jorm AF. White matter hyperintensities are related to physical disability and poor motor function. J Neurol Neurosurg Psychiatry. 2005 Mar;76(3):362–7

24. Kloppenborg RP, Nederkoorn PJ, Geerlings MI, van den Berg E. Presence and progression of white matter hyperintensities and cognition: a meta-analysis. Neurology. 2014 Jun 10;82(23):2127–38

25. Grangeon MC, Seixas C, Quarantini LC, Miranda-Scippa A, Pompili M, Steffens DC, Wenzel A, Lacerda AL, de Oliveira IR. White matter hyperintensities and their association with suicidality in major affective disorders: a meta-analysis of magnetic resonance imaging studies. CNS Spectr. 2010 Jun;15(6):375–81.

26. Mayer C, Frey BM, Schlemm E, Petersen M, Engelke K, Hanning U, Jagodzinski A, Borof K, Fiehler J, Gerloff C, Thomalla G, Cheng B. Linking cortical atrophy to white matter hyperintensities of presumed vascular origin. J Cereb Blood Flow Metab. 2021 Jul;41(7):1682–1691.

27. Hofman A, Breteler MM, van Duijn CM, Krestin GP, Pols HA, Stricker BH, Tiemeier H, Uitterlinden AG, Vingerling JR, Witteman JC. The Rotterdam Study: objectives and design update. Eur J Epidemiol. 2007;22(11):819–29.

28. Blom JW, de Ruijter W, Witteman JC, Assendelft WJ, Breteler MM, Hofman A, Gussekloo J. Changing prediction of mortality by systolic blood pressure with increasing age: the Rotterdam study. Age (Dordr). 2013 Apr;35(2):431–8.

29. Ikram MA, van der Lugt A, Niessen WJ, Koudstaal PJ, Krestin GP, Hofman A, Bos D, Vernooij MW. The Rotterdam Scan Study: design update 2016 and main findings. Eur J Epidemiol. 2015 Dec;30(12):1299–315.

30. Fischl B. FreeSurfer. Neuroimage. 2012 Aug 15;62(2):774–81.

31. Rorden C, Karnath HO, Bonilha L. MRIcron dicom to nifti converter. Neuroimaging Informatics Tools and Resources Clearinghouse (NITRC). 2012.

32. Jenkinson, M, Beckmann, CF, Behrens, TEJ, Woolrich, MW, & Smith, SM. FSL (FMRIB Software Library). Neuroimage. 2012;62(2), 782–790.

33. de Groot M, Ikram MA, Akoudad S, Krestin GP, Hofman A, van der Lugt A, Niessen WJ, Vernooij MW. Tract-specific white matter degeneration in aging: the Rotterdam Study. Alzheimers Dement. 2015 Mar;11(3):321–30.

34. Park G, Hong J, Duffy BA, Lee JM, Kim H. White matter hyperintensities segmentation using the ensemble U-Net with multi-scale highlighting foregrounds. Neuroimage. 2021;237:118140.

35. Harrell Jr FE. Cox proportional hazards regression model. In: Regression Modeling Strategies: With Applications to Linear Models, Logistic Regression, and Survival Analysis 2001 (pp. 465–507). New York, NY: Springer New York.

36. VanderWeele TJ. Causal mediation analysis with survival data. Epidemiology. 2011 Jul;22(4):582–5

37. Benjamini, Y. and Hochberg, Y. (1995), Controlling the False Discovery Rate: A Practical and Powerful Approach to Multiple Testing. Journal of the Royal Statistical Society: Series B (Methodological), 57: 289–300.

38. Maillard P, Carmichael O, Harvey D, Fletcher E, Reed B, Mungas D, DeCarli C. FLAIR and diffusion MRI signals are independent predictors of white matter hyperintensities. AJNR Am J Neuroradiol. 2013 Jan;34(1):54–61.

39. van Leijsen EMC, Kuiperij HB, Kersten I, Bergkamp MI, van Uden IWM, Vanderstichele H, Stoops E, Claassen JAHR, van Dijk EJ, de Leeuw FE, Verbeek MM. Plasma Aβ (Amyloid-β) Levels and Severity and Progression of Small Vessel Disease. Stroke. 2018 Apr;49(4):884–890..

40. Mayer C, Frey BM, Schlemm E, Petersen M, Engelke K, Hanning U, Jagodzinski A, Borof K, Fiehler J, Gerloff C, Thomalla G, Cheng B. Linking cortical atrophy to white matter hyperintensities of presumed vascular origin. J Cereb Blood Flow Metab. 2021 Jul;41(7):1682–1691.

41. Tuladhar AM, Reid AT, Shumskaya E, de Laat KF, van Norden AG, van Dijk EJ, Norris DG, de Leeuw FE. Relationship between white matter hyperintensities, cortical thickness, and cognition. Stroke. 2015 Feb;46(2):425–32

42. Kuang Q, Huang M, Lei Y, Wu L, Jin C, Dai J, Zhou F. Clinical and cognitive correlates tractography analysis in patients with white matter hyperintensity of vascular origin. Front Neurosci. 2023 Jun 16;17:1187979.

43. Ab Aziz CB, Ahmad AH. The role of the thalamus in modulating pain. Malays J Med Sci. 2006;13(2):11–8.

44. K Dharani. Functional anatomy of the brain. In: The biology of thought: A neuronal mechanism in the generation of thought – A new molecular model (pp. 3–29). Elsevier. 2015

45. Stankewitz A, Mayr A, Irving S, Witkovsky V, Schulz E. Pain and the emotional brain: pain-related cortical processes are better reflected by affective evaluation than by cognitive evaluation. Sci Rep. 2023 May 22;13(1):8273.

46. Yoshino A, Okamoto Y, Doi M, Okada G, Takamura M, Ichikawa N, Yamawaki S. Functional Alterations of Postcentral Gyrus Modulated by Angry Facial Expressions during Intraoral Tactile Stimuli in Patients with Burning Mouth Syndrome: A Functional Magnetic Resonance Imaging Study. Front Psychiatry. 2017 Nov 6;8:224.

47. Mayr A, Jahn P, Stankewitz A, Deak B, Winkler A, Witkovsky V, Eren O, Straube A, Schulz E. Patients with chronic pain exhibit individually unique cortical signatures of pain encoding. Hum Brain Mapp. 2022 Apr 1;43(5):1676–1693.

48. Emerson NM, Zeidan F, Lobanov OV, Hadsel MS, Martucci KT, Quevedo AS, Starr CJ, Nahman-Averbuch H, Weissman-Fogel I, Granovsky Y, Yarnitsky D, Coghill RC. Pain sensitivity is inversely related to regional grey matter density in the brain. Pain. 2014 Mar;155(3):566–573.

49. Grant JA, Courtemanche J, Duerden EG, Duncan GH, Rainville P. Cortical thickness and pain sensitivity in zen meditators. Emotion. 2010 Feb;10(1):43–53.

50. Villemure C, Ceko M, Cotton VA, Bushnell MC. Insular cortex mediates increased pain tolerance in yoga practitioners. Cereb Cortex. 2014 Oct;24(10):2732–40.

51. Kramer JLK, Jutzeler CR, Haefeli J, Curt A, Freund P. Discrepancy between perceived pain and cortical processing: A voxel-based morphometry and contact heat evoked potential study. Clin Neurophysiol. 2016 Jan;127(1):762–768.

52. Ruscheweyh R, Wersching H, Kugel H, Sundermann B, Teuber A. Gray matter correlates of pressure pain thresholds and self-rated pain sensitivity: a voxel-based morphometry study. Pain. 2018 Jul;159(7):1359–1365

53. de Kruijf M, Bos D, Huygen FJ, Niessen WJ, Tiemeier H, Hofman A, Uitterlinden AG, Vernooij MW, Ikram MA, van Meurs JB. Structural Brain Alterations in Community Dwelling Individuals with Chronic Joint Pain. AJNR Am J Neuroradiol. 2016 Mar;37(3):430–8.

54. Neumann L, Wulms N, Witte V, Spisak T, Zunhammer M, Bingel U, Schmidt-Wilcke T. Network properties and regional brain morphology of the insular cortex correlate with individual pain thresholds. Hum Brain Mapp. 2021 Oct 15;42(15):4896–4908.

55. Schwedt TJ, Chong CD. Correlations between brain cortical thickness and cutaneous pain thresholds are atypical in adults with migraine. PLoS One. 2014 Jun 16;9(6):e99791

56. Niddam DM, Wang SJ, Tsai SY. Pain sensitivity and the primary sensorimotor cortices: a multimodal neuroimaging study. Pain. 2021 Mar 1;162(3):846–855.

57. Schlereth T, Baumgärtner U, Magerl W, Stoeter P, Treede RD. Left-hemisphere dominance in early nociceptive processing in the human parasylvian cortex. Neuroimage. 2003 Sep;20(1):441–54.

58. Jin SH, Lee SH, Yang ST, An J. Hemispheric asymmetry in hand preference of right-handers for passive vibrotactile perception: an fNIRS study. Sci Rep. 2020 Aug 7;10(1):13423.

59. Jacobsen BK, Eggen AE, Mathiesen EB, Wilsgaard T, Njolstad I. Cohort profile: the tromso study. Int J Epidemiol 2012;41:961–7.

60. Ikram MA, Kieboom BCT, Brouwer WP, Brusselle G, Chaker L, Ghanbari M, Goedegebure A, Ikram MK, Kavousi M, de Knegt RJ, Luik AI, van Meurs J, Pardo LM, Rivadeneira F, van Rooij FJA, Vernooij MW, Voortman T, Terzikhan N. The Rotterdam Study. Design update and major findings between 2020 and 2024. Eur J Epidemiol. 2024;39(2):183–206.

61. Dostrovsky JO. Role of thalamus in pain. Prog Brain Res. 2000;129:245–57.

62. Wolff M, Vann SD. The Cognitive Thalamus as a Gateway to Mental Representations. J Neurosci. 2019;39(1):3–14.

63. Wardlaw JM, Valdés Hernández MC, Muñoz-Maniega S. What are white matter hyperintensities made of? Relevance to vascular cognitive impairment. J Am Heart Assoc. 2015 Jun 23;4(6):001140.

