## Supplementary for "Structural Brain Pathways Linking White Matter Hyperintensities to Pain Sensitivity"

**Table S1: Global-level analysis between brain measurements and CPT.** Covariates: age, sex, and ICV. The Rotterdam Study.

| **Brain measurements** | **Hazards ratio** | **95% CI** | ***p*-value** | **FDR-*p*** |
| --- | --- | --- | --- | --- |
| Total brain volume | 0.92 | 0.81-1.04 | 0.177 | 0.226 |
| Total brain volume (left-hemisphere) | 0.92 | 0.81-1.04 | 0.188 | 0.226 |
| Total brain volume (right-hemisphere) | 0.92 | 0.81-1.04 | 0.177 | 0.226 |
| Total gray matter | 0.92 | 0.82-1.02 | 0.126 | 0.226 |
| Total gray matter (left-hemisphere) | 0.92 | 0.82-1.02 | 0.122 | 0.226 |
| Total gray matter (right-hemisphere) | 0.92 | 0.83-1.03 | 0.140 | 0.226 |
| Total white matter | 0.96 | 0.86-1.07 | 0.480 | 0.508 |
| Total white matter (left-hemisphere) | 0.97 | 0.87-1.08 | 0.525 | 0.525 |
| Total white matter (right-hemisphere) | 0.96 | 0.86-1.07 | 0.446 | 0.502 |
| Total WMH | 1.06 | 1.00-1.12 | 0.061 | 0.226 |
| Total WMH (left-hemisphere) | 1.06 | 1.00-1.13 | 0.047* | 0.226 |
| Total WMH (right-hemisphere) | 1.06 | 1.00-1.12 | 0.067 | 0.226 |
| Mean FA | 0.94 | 0.87-1.01 | 0.085 | 0.226 |
| Mean FA (left-hemisphere) | 0.92 | 0.85-1.00 | 0.050 | 0.226 |
| Mean FA (right-hemisphere) | 0.95 | 0.89-1.02 | 0.144 | 0.226 |
| Mean MD | 1.06 | 0.98-1.15 | 0.134 | 0.226 |
| Mean MD (left-hemisphere) | 1.05 | 0.98-1.13 | 0.151 | 0.226 |
| Mean MD (right-hemisphere) | 1.06 | 0.98-1.16 | 0.137 | 0.226 |

WMH: Log-transformed white matter hyperintensity; FA: fractional anisotropy; MD: mean diffusivity;

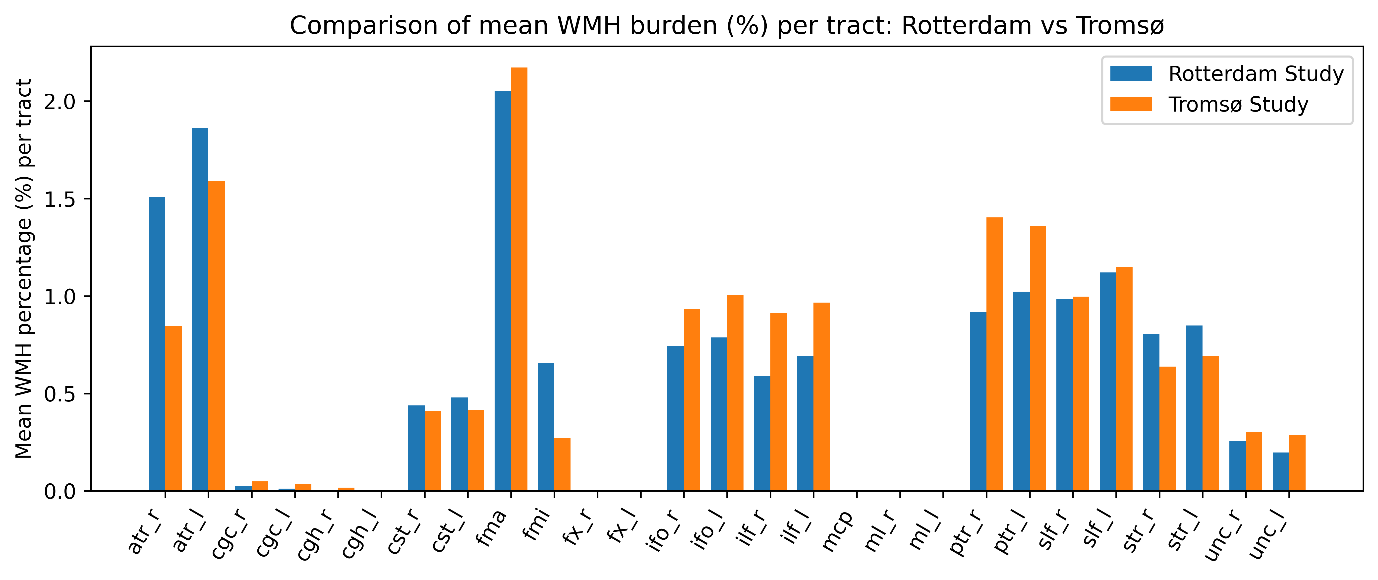

**Figure S1: Distribution of WMH burden across the 27 predefined white matter tracts in the Rotterdam Study and the Tromsø Study.** Each bar represents the WMH percentage relative to the total volume of the corresponding tract (100% indicating complete tract coverage). Abbreviations: WMH : white matter hyperintensities; FA: fractional anisotropy; MD: mean diffusivity; ATR, anterior thalamic radiation; IFO, inferior fronto-occipital fasciculus; ILF, inferior longitudinal fasciculus; PTR, posterior thalamic radiation; SLF, superior longitudinal fasciculus; UNC, uncinate fasciculus; FMA, forceps major; FMI, forceps minor; CGC, cingulate gyrus part of cingulum; CGH, parahippocampal part of cingulum; CST, corticospinal tract; MCP, middle cerebellar peduncle; ML, medial lemniscus; STR, superior thalamic radiation; FX, fornix.

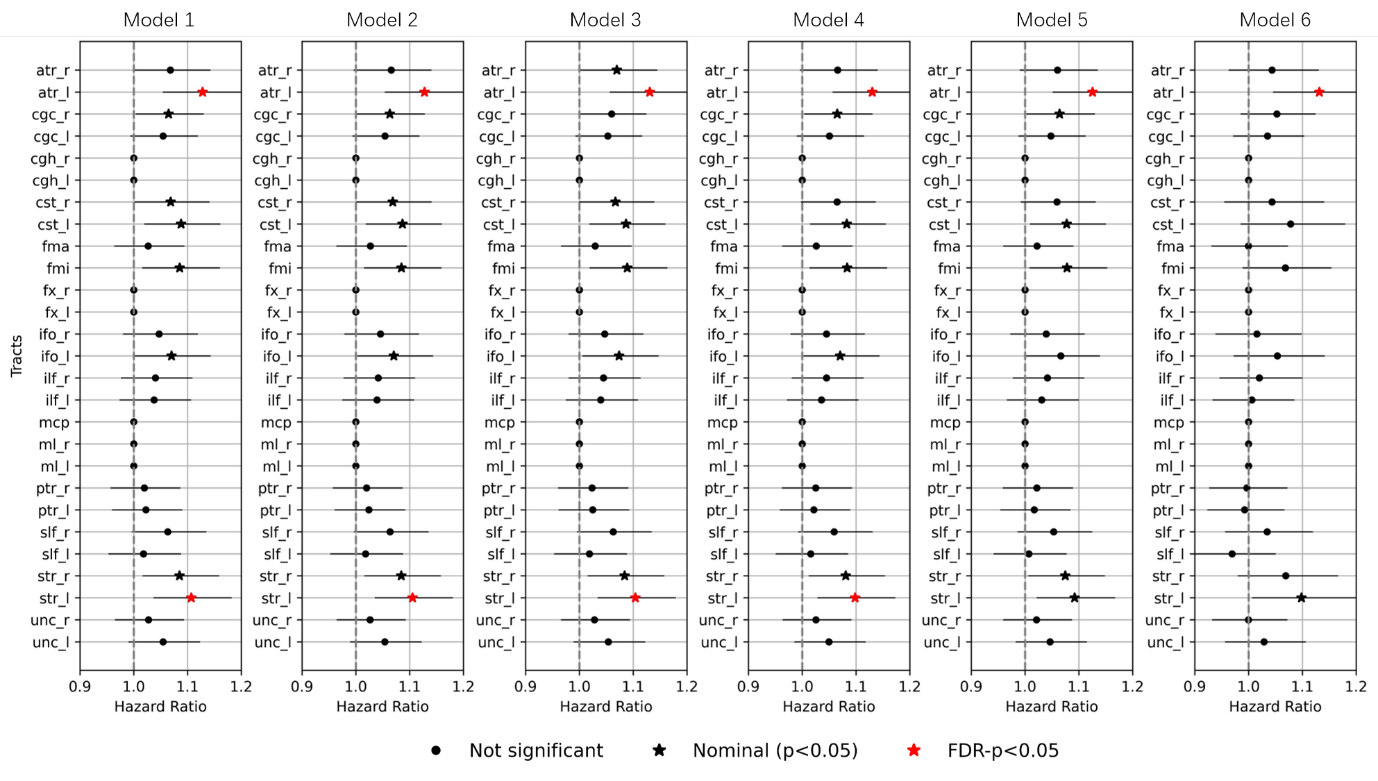

**Figure S2: Sensitivity analysis of Cox regression between cold pressor test (CPT) performance and tract-specific WMH in the discovery cohort under sequential covariate adjustment.** Model 1 included age, sex, total WMH volume, and intracranial volume (ICV). Models 2–6 further adjusted sequentially as follows: Model 2 = Model 1 + chronic pain; Model 3 = Model 2 + educational years; Model 4 = Model 3 + pain medication use; Model 5 = Model 4 + hypertension; Model 6 = Model 5 + smoking. WMH was log-transformed and normalized to a 0–1 scale in the analysis. Abbreviations: WMH : white matter hyperintensities; ATR, anterior thalamic radiation; IFO, inferior fronto-occipital fasciculus; ILF, inferior longitudinal fasciculus; PTR, posterior thalamic radiation; SLF, superior longitudinal fasciculus; UNC, uncinate fasciculus; FMA, forceps major; FMI, forceps minor; CGC, cingulate gyrus part of cingulum; CGH, parahippocampal part of cingulum; CST, corticospinal tract; MCP, middle cerebellar peduncle; ML, medial lemniscus; STR, superior thalamic radiation; FX, fornix. _L and _R denote left and right hemispheres, respectively. FDR: false discovery rate. The Rotterdam Study.

**Table S2A: Mediation analysis using tract-connected cortical regions as potential mediators, in the Rotterdam Study.** Exposure variable: log-transformed WMH volume within a tract; Outcome variable: CPT, analysed as a time-to-event measure of pain sensitivity

| **Tract name -> connected cortical region** | **Proportion of mediation**  **(%, *p*-value)** | **Total effect**  **(*p*-value)** | **ADE (direct)**  **(*p*-value)** | **ACME (indirect)**  **(*p*-value)** |
| --- | --- | --- | --- | --- |
| CST_L -> Postcentral_L (Volume) | 5.5%, 0.044* | 0.014* | 0.018* | 0.030* |
| CST_L -> Precentral_L (Volume) | 5.4%, 0.048* | 0.014* | 0.018* | 0.034* |
| ATR_R -> Pars orbitalis_R (Volume) | 6.8%, 0.072 | 0.032* | 0.054 | 0.040* |
| STR_L -> Postcentral_L (Volume) | 4.3%, 0.046* | <0.001* | <0.001* | 0.046* |
| STR_L -> Precentral_L (Volume) | 3.1%, 0.048* | <0.001* | 0.002* | 0.048* |
| SLF_R Postcentral_R (Thickness) | 5.8%, 0.156 | 0.072 | 0.054 | 0.084 |
| ATR_L -> Pars triangularis_L (Thickness) | 2.9%, 0.090 | <0.001* | <0.001* | 0.090 |
| ATR_R -> Rostral middle frontal_R (Thickness) | 7.7%, 0.104 | 0.018* | 0.064 | 0.090 |
| FMI -> Rostral middle frontal_R (Thickness) | 6.8%, 0.098 | <0.001* | 0.006* | 0.098 |
| IFO_L -> Middle temporal_L (Volume) | 4.8%, 0.118 | 0.014* | 0.034* | 0.108 |
| ILF_L -> Inferior temporal_L (Volume) | 7.7%, 0.392 | 0.300 | 0.368 | 0.132 |
| UNC_L -> Medial orbitofrontal_L (Volume) | 4.2%, 0.196 | 0.052 | 0.042* | 0.144 |
| UNC_L -> Middle temporal_L (Volume) | 3.7%, 0.186 | 0.044* | 0.058 | 0.146 |
| ATR_R -> Pars opercularis_R (Thickness) | 3.9%, 0.204 | 0.050 | 0.062 | 0.162 |
| ATR_R -> Pars orbitalis_R (Thickness) | 6.8%, 0.226 | 0.048* | 0.066 | 0.186 |
| FMA -> Pericalcarine_L (Volume) | 6.9%, 0.632 | 0.572 | 0.490 | 0.188 |
| ILF_R -> Middle temporal_R (Volume) | 5.1%, 0.478 | 0.366 | 0.406 | 0.188 |
| UNC_R -> Middle temporal_R (Volume) | 3.5%, 0.438 | 0.304 | 0.332 | 0.190 |
| UNC_L -> Inferior temporal_L (Volume) | 5.5%, 0.260 | 0.070 | 0.084 | 0.194 |
| ILF_L -> Middle temporal_L (Volume) | 3.9%, 0.452 | 0.328 | 0.366 | 0.200 |
| … | … | … | … | … |

ACME: Average causal mediated effect; ADE: Average direct effect;

ATR, anterior thalamic radiation; IFO, inferior fronto-occipital fasciculus; ILF, inferior longitudinal fasciculus; PTR, posterior thalamic radiation; SLF, superior longitudinal fasciculus; UNC, uncinate fasciculus; FMA, forceps major; FMI, forceps minor; CGC, cingulate gyrus part of cingulum; CGH, parahippocampal part of cingulum; CST, corticospinal tract; MCP, middle cerebe llar peduncle; ML, medial lemniscus; STR, superior thalamic radiation; FX, fornix.

_L and _R denote left and right hemispheres, respectively; * indicates statistical significance (e.g., p < 0.05).

Covariates included age, sex, total WMH volume, and intracranial volume (ICV).

Results are presented in ascending order of the ACME p-value.

The completed table can be found online:

<https://drive.google.com/file/d/1jiFx7ZC7oYZJrYTpTw_9_gYoi53sc_or/view?usp=sharing>

**Table S2B: Mediation analysis using tract-connected cortical regions as potential mediators, in the Tromsø Study.** Exposure variable: log-transformed WMH volume within a tract; Outcome variable: CPT, analysed as a time-to-event measure of pain sensitivity.

| **Tract name -> connected cortical region** | **Proportion of mediation**  **(%, *p*-value)** | **Total effect**  **(*p*-value)** | **ADE (direct)**  **(*p*-value)** | **ACME (indirect)**  **(*p*-value)** |
| --- | --- | --- | --- | --- |
| CST_L -> Postcentral_L (Volume) | 4.7%, 0.866 | 0.844 | 0.730 | 0.042* |
| CST_L -> Precentral_L (Volume) | 10.5%, 0.860 | 0.850 | 0.684 | 0.026* |
| ATR_R -> Pars orbitalis_R (Volume) | 0.2%, 0.930 | 0.344 | 0.340 | 0.922 |
| STR_L -> Postcentral_L (Volume) | 8.2%, 0.798 | 0.790 | 0.918 | 0.032* |
| STR_L -> Precentral_L (Volume) | 11.3%, 0.798 | 0.790 | 0.958 | 0.028* |
| SLF_R -> Postcentral_R (Thickness) | 0.8%, 0.804 | 0.464 | 0.468 | 0.676 |
| ATR_L -> Pars triangularis_L (Thickness) | 9.1%, 0.460 | 0.402 | 0.492 | 0.090 |
| ATR_R -> Rostral middle frontal_R (Thickness) | 0.2%, 0.932 | 0.344 | 0.340 | 0.916 |
| FMI -> Rostral middle frontal_R (Thickness) | 0.0%, 0.934 | 0.030* | 0.028* | 0.932 |
| IFO_L -> Middle temporal_L (Volume) | 5.4%, 0.510 | 0.466 | 0.526 | 0.116 |
| ILF_L -> Inferior temporal_L (Volume) | 5.0%, 0.520 | 0.436 | 0.496 | 0.172 |
| UNC_L -> Medial orbitofrontal_L (Volume) | 0.0%, 0.964 | 0.248 | 0.250 | 0.916 |
| UNC_L -> Middle temporal_L (Volume) | 10.7%, 0.320 | 0.304 | 0.396 | 0.024* |
| ATR_R -> Pars opercularis_R (Thickness) | 2.1%, 0.708 | 0.332 | 0.364 | 0.596 |
| ATR_R -> Pars orbitalis_R (Thickness) | 0.3%, 0.960 | 0.314 | 0.316 | 0.902 |
| FMA -> Pericalcarine_L (Volume) | 3.2%, 0.556 | 0.126 | 0.120 | 0.518 |
| ILF_R -> Middle temporal_R (Volume) | 0.7%, 0.690 | 0.024* | 0.020* | 0.682 |
| UNC_R -> Middle temporal_R (Volume) | 1.0%, 0.858 | 0.734 | 0.770 | 0.396 |
| UNC_L -> Inferior temporal_L (Volume) | 13.1%, 0.318 | 0.296 | 0.402 | 0.026* |
| ILF_L -> Middle temporal_L (Volume) | 6.5%, 0.506 | 0.470 | 0.544 | 0.056 |
| … | … | … | … | … |

ACME: Average causal mediated effect; ADE: Average direct effect;

ATR, anterior thalamic radiation; IFO, inferior fronto-occipital fasciculus; ILF, inferior longitudinal fasciculus; PTR, posterior thalamic radiation; SLF, superior longitudinal fasciculus; UNC, uncinate fasciculus; FMA, forceps major; FMI, forceps minor; CGC, cingulate gyrus part of cingulum; CGH, parahippocampal part of cingulum; CST, corticospinal tract; MCP, middle cerebe llar peduncle; ML, medial lemniscus; STR, superior thalamic radiation; FX, fornix.

_L and _R denote left and right hemispheres, respectively; * indicates statistical significance (e.g., p < 0.05).

Covariates included age, sex, total WMH volume, and intracranial volume (ICV).

Results are presented in the same order as in Supplementary Table S2A.

The completed table can be found online:

<https://drive.google.com/file/d/1Rv_bpOmdFSj3PNw-VZJOBy28q50IYRpq/view?usp=sharing>
